# Genomewide association study of epiretinal membrane: discovery of significant risk loci in each of three American populations

**DOI:** 10.1101/2023.04.25.23289093

**Authors:** Joel Gelernter, Daniel Levey, Marco Galimberti, Kelly Harrington, Hang Zhou, Keyrun Adhikari, J. Michael Gaziano, Dean Eliott, Murray B. Stein

**Affiliations:** Department of Psychiatry, Yale School of Medicine, New Haven, CT; Department of Psychiatry, VA Connecticut Healthcare Center, West Haven, CT; Departments of Genetics and Neuroscience, Yale School of Medicine, New Haven, CT; Massachusetts Veterans Epidemiology Research and Information Center (MAVERIC), VA Boston Healthcare System, Boston, MA; Department of Psychiatry, Boston University Chobanian & Avedisian School of Medicine,, Boston, MA; Department of Medicine, Harvard Medical School, Boston, MA; Department of Medicine, Divisions of Aging and Preventative Medicine, Brigham and Women’s Hospital, Boston, MA; Massachusetts Eye and Ear, Department of Ophthalmology, Harvard Medical School, Boston, MA; University of California, San Diego, La Jolla, CA; VA San Diego Healthcare System, San Diego, CA

**Keywords:** Million Veteran Program (MVP), genomewide association study (GWAS), epiretinal membrane (ERM), macular pucker, multi-ancestry study, transcriptome wide association study (TWAS), pleiotropy

## Abstract

**IMPORTANCE:** Epiretinal membrane (ERM) is a common retinal condition characterized by the presence of fibrocellular tissue on the retinal surface, often with consequent loss of vision and visual distortion.

**OBJECTIVE:** Genomewide association studies (GWAS) can reveal the biology underlying complex genetic traits like ERM; there have been no previous large-scale GWAS of this trait.

**DESIGN:** We used electronic health record diagnosis to identify Million Veteran Program (MVP) participants with ERM in three populations for genomewide association analysis and further statistical investigation of the results.

**SETTING:** Veterans who volunteered for the nationwide Department of Veterans Affairs MVP study, eligible because they have used Veterans Health Administration facilities

**PARTICIPANTS:** 31,374 European-American (EUR) cases and 414,052 EUR controls, 4,739 African-American (AFR) cases and 107,773 AFR controls, and 2,119 Latino (Admixed American, AMR) cases and 36,163 AMR controls – a total of 38,232 cases and 557,988 controls.

**METHODS:** We completed GWAS in each population separately, then results were meta-analyzed. We also evaluated genetic correlation with other traits in external samples, and completed pathway enrichment analyses.

**MAIN OUTCOME MEASURES:** Genomewide-significant association with ERM.

**RESULTS:** Genomewide significant associations were observed in all three populations studied: 31 risk loci in EUR subjects, 3 in AFR, and 2 in AMR, with 48 identified in trans-ancestry meta-analysis. The most strongly associated locus in both EUR (rs9823832, p=9.06×10^−37^) and the meta-analysis (rs28630834, p=2.90×10^−37^) was *DHX36* (DEAH-Box Helicase 36). We investigated expression quantitative trait locus associations for eye related function and found several GWS variants associate to alterations in gene expression in the macula, including *DHX36*\*rs9438. ERM showed significant genetic correlation to depression and to disorders of the vitreous. Pathway enrichment analyses implicated collagen and collagen-adjacent mechanisms, among others.

**CONCLUSIONS AND RELEVANCE:** This well-powered ERM GWAS has identified novel genetic associations, some very strong, that point to biological mechanisms for ERM and merit further investigation.

## Introduction

Epiretinal membrane (ERM), sometimes called macular pucker, is characterized by the proliferation of cells on the retinal surface. These cells adhere to the retina, produce extracellular matrix, and develop contractile properties, resulting in tangential retinal traction and distortion of the retinal architecture. Symptoms can be severe and require surgery; patients may complain of loss of central vision and/or visual distortion (metamorphopsia, “wavy vision”). Diagnosis can be established by ophthalmoscopy, fundus photography, and optical coherence tomography (OCT). It is common, with prevalence of 2% in patients age >50 and 20% age >75. Age is associated with increased risk, and smoking may be associated with decreased risk^1^. The most common cause is posterior vitreous detachment, an age-related separation of the vitreous from the retina. Most patients do not require intervention; for those with symptomatic ERM, vitrectomy with epiretinal membrane peeling is the typical treatment and results are variable; there is no nonsurgical treatment. While many of the pathophysiological processes that lead to ERM are known and there is considerable knowledge of its biology^2^, the underlying molecular genetic risk factors are not understood.

Genomewide association studies (GWAS) are presently the most useful, and most universally applicable, tools to understand the biology underlying genetically complex traits such as ERM. Large GWAS have been previously performed to identify common variant associations with defects related to the macula, but these have focused mostly on age-related macular degeneration (AMD)^3^ (including the first major GWAS^4^.) A GWAS study in the UK Biobank considered AMD, diabetic retinopathy, retinal detachment, glaucoma, and myopia, with risk loci mapped for each trait and pleiotropy between them^5^. A large GWAS of cataract in UK Biobank^6^, Genetic Epidemiology Research on Adult Health and Aging (GERA)^7^, and 23andme (a personal genomics company) reported 54 genomewide significant (GWS) risk loci. ERM, though, has, to our knowledge, not been studied with respect to genetics using any genomewide approach.

To identify a large number of risk genes, novel biology, and phenotypic subtypes, GWAS with large sample size are necessary. The Department of Veterans Affairs (VA) Million Veteran Program (MVP) is one of the world’s largest biobanks including genetic, environmental, and medical information, based on data from United States military veterans.^8-11^. The MVP has enrolled >900,000 participants, with excellent representation from non-European-ancestry participants. Genotype data are currently available for >650,000 participants. The MVP sample is relatively old (55% are between ages 50-69^10^) and ill – subjects are enrolled based on their use of the US Veterans Health Administration system. We conducted a GWAS of ERM in the MVP sample, using diagnoses based on the electronic health record (EHR).

## Subjects and Methods

### MVP: Primary analyses

We used data release version 4 of the MVP.^12^ Linked and de-identified EHRs were queried using the Veterans Affairs Informatics and Computing Infrastructure to identify individuals with International Classification of Disease (ICD) codes for ERM, specified in the Supplementary Methods. These codes identified 31,374 European-American (EUR) cases and 414,052 EUR controls, 4,739 African-American (AFR) cases and 107,773 AFR controls, and 2,119 AMR (Admixed American or Latino) cases and 36,163 AMR controls; more detail is provided in Table 1. Research involving the MVP in general is approved by the VA Central Institutional Review Board. All participants provided written informed consent.

**Table 1:** MVP sample demographic information.

| EUR | Case | Control | Total |
| --- | --- | --- | --- |
| Male | 30176 | 382566 | 412742 |
| Age (SEM) | 71.15(0.05) | 64.22(0.02) |  |
| Female | 1198 | 31486 | 32684 |
| Age (SEM) | 64.78(0.78) | 52.10(0.08) |  |
| Total (EUR) | 31374 | 414052 | 445426 |
| AFR | Case | Control | Total |
| Male | 4398 | 92592 | 96990 |
| Age (SEM) | 66.63(0.13) | 58.66(0.04) |  |
| Female | 341 | 15181 | 15522 |
| Age (SEM) | 59.59(0.51) | 48.75(0.09) |  |
| Total (AFR) | 4739 | 107773 | 112512 |
| AMR | Case | Control | Total |
| Male | 2055 | 32611 | 34666 |
| Age (SEM) | 68.25(0.20) | 55.44(0.09) |  |
| Female | 64 | 3552 | 3616 |
| Age (SEM) | 62.4(0.91) | 43.29(0.218) |  |
| Total (AMR) | 2119 | 36163 | 38282 |
| TOTAL (All) | 38232 | 557988 | 596220 |

### FinnGen: Phenotypes for pleiotropy analyses

We accessed GWAS summary statistics from FinnGen^13^, a Finnish nationwide biobank collection, data freeze version 7, for two traits potentially relating to ERM, “diseases of the vitreous body” and “glaucoma”. ICD-10 codes used for diagnosis definition, Supplementary Methods.

#### Genotyping, Imputation, Quality Control (QC), and GWAS

Genotyping and imputation of MVP subjects has been described previously.^10,14^ Briefly, a customized Affymetrix Axiom Array was used for genotyping. MVP genotype data for biallelic SNPs were imputed using Minimac4 and a reference panel from the African Genome Resources (AGR) by the Sanger Institute. Details, Supplement.

For the FinnGen dataset^13^, results were downloaded from the FinnGen website after registration (https://www.finngen.fi/en/access_results). Analyses details are given in the Supplement.

#### Heritability and Genetic correlations

Linkage disequilibrium score regression (LDSC)^15^ was used to calculate liability scale SNP-heritability for EUR, AFR and AMR ancestry of MVP data for ERM. We used a value of 0.091 for the population prevalence ^1^. For EUR, this was calculated directly with LDSC. To estimate liability scale SNP-heritability for AFR and AMR cohorts, we calculated LD scores with cov-LDSC ^16^ from 10,000 random independent individuals for the SNPs identified in the HapMap Project ^17^. LDSC was also used to calculate genetic correlation^18^ between the EUR MVP data for ERM and four other traits: two ophthalmologic traits, glaucoma^19^ and disorders of the vitreous body^13^; and two psychiatric traits, posttraumatic stress disorder (PTSD)^20^ and depression^21^.

#### Cross-ancestry Fine-mapping

We did fine-mapping leveraging LD information from multiple ancestries to identify potential causal variants using MsCAVIAR^22^. Details, Supplement.

#### Expression QTLs (eQTL)

GWAS significant loci were investigated to determine whether they were eQTLs using an eye tissue specific database^*23*^. Lead variants from the EUR ancestry GWAS were looked up first; if lead variants were not available proxy SNPs were selected from genomewide significant (GWS) variants in the same locus.

#### Transcriptome-Wide Association Study (TWAS) and Fine-mapping

We performed TWAS using FUSION^24^ for the EUR GWAS statistics. Details, Supplement.

#### MAGMA gene-based and gene set analyses

We used MAGMA ^25^ implemented in FUMA ^26^ for gene-based and gene-set analyses. Details, Supplement.

## Results

### MVP analyses

GWS associations were observed in all three populations studied. 31 risk loci were identified in EUR subjects, 3 in AFR, and 2 in AMR, reflecting the greater power of the analysis in EUR compared to the other two populations presumably due to difference in sample sizes (Table 2a-c; Figures S1-S3).

**Table 2:**
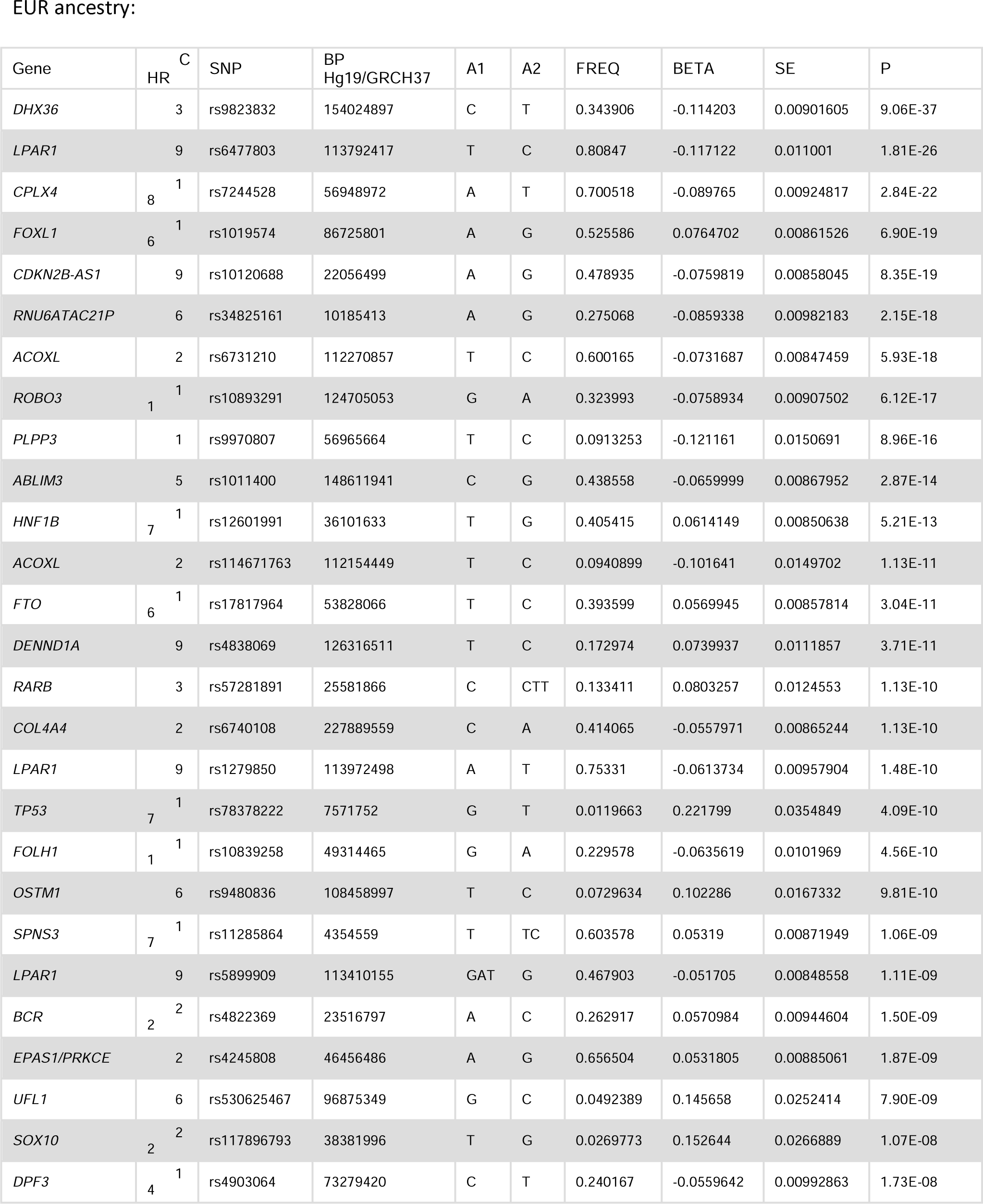
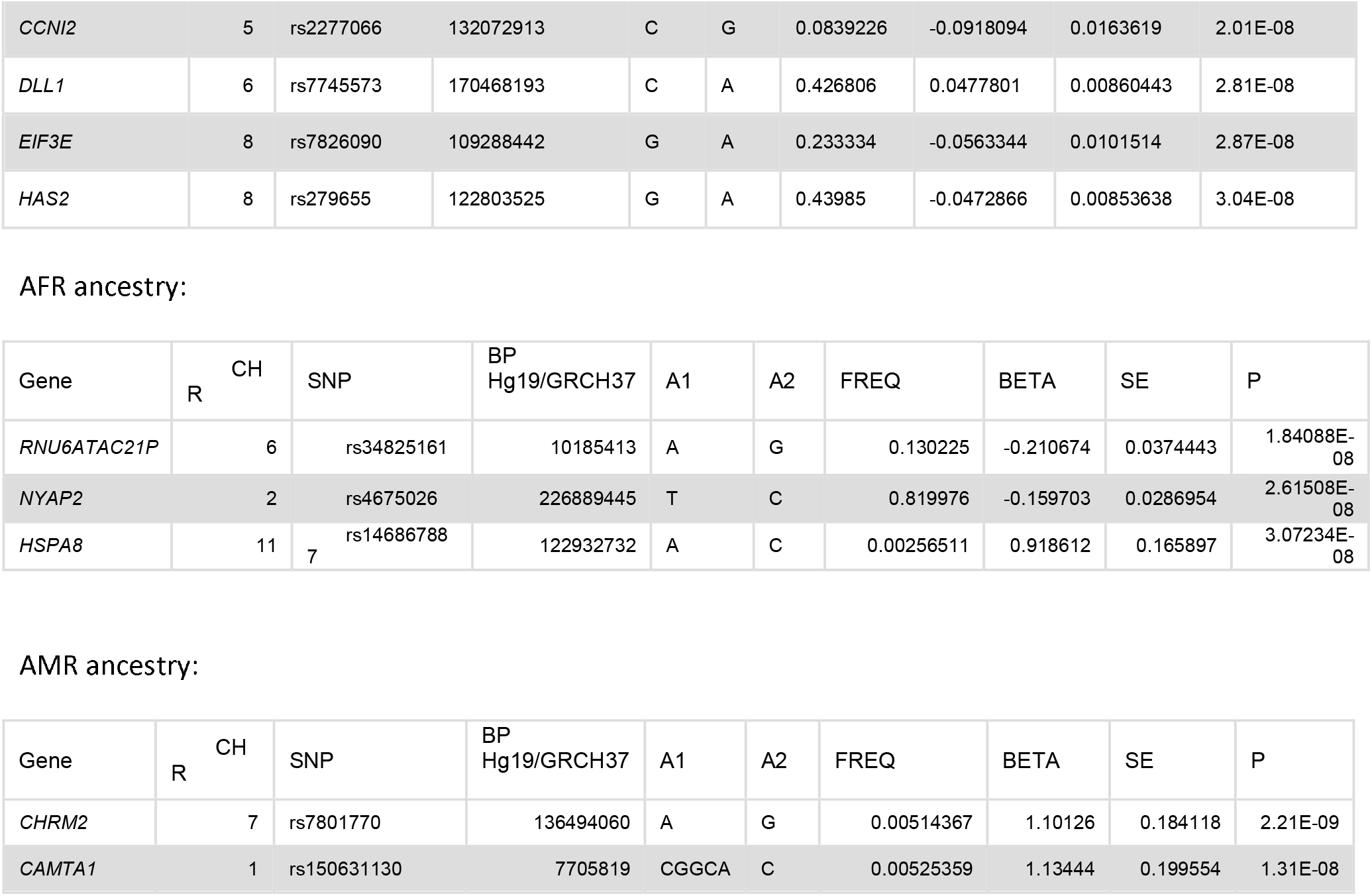
Independent significant risk loci identified in EUR, AFR, and AMR. A1, risk allele; FREQ, frequency of A1 (sorted by p-value)

In EUR, of the 31 GWS loci, 14 were associated with a p-value <10^−10^. The most strongly supported variant was rs9823832 (p=9.06×10^−37^) which maps to *DHX36* (DEAH-Box Helicase 36). Other associated variants highlighted for biological interest (Discussion) include *CPLX4* (rs7244528 associated at 2.84×10^−22^), *ROBO3*, to which variant rs10893291 maps (associated at 6.12×10^−17^), and rs1011400 at locus *ABLIM3* (p=2.87×10^−14^). Additional associated loci include *FTO*\*rs17817964 (p=3.04×10^−11^) (FTO Alpha-Ketoglutarate Dependent Dioxygenase) and *RARB*\*rs57281891 (p=1.13×10^−10^) (Retinoic Acid Receptor Beta). Other associated variants map to *CDKN2B-AS1* (rs10120688, p=8.35×10-19) and *ACOXL* – two independent variants >115,000 basepairs apart, rs6731210 (p=5.93×10^−18^) and rs114671763 (p=1.13×10^−11^). In AMR, the most strongly associated locus (p=2.21×10^−9^) was *CHRM2*\*rs7801770, which encodes Cholinergic Receptor Muscarinic 2. (This *CHRM2* SNP is not polymorphic in EUR and non-significant in AFR (p=0.44).)

The trans-ancestry meta-analysis identified 48 GWS variants (Figure 1, Table 3); many were not significant (at p≤5×10^−8^) in any of the population-specific analyses. Of these 48 variants, 27 regions had p-value stronger than 10^−10^. In numerous cases, the same gene was implicated in the meta-analysis as for EUR but with a different lead variant; the same variant as for EUR-only and meta-analysis was implicated in 15 instances (Table 3). In the trans-ancestry meta-analysis, the most strongly supported variant maps to *DHX36* as for EUR – a different variant at the same risk locus, rs28630834 (p=2.90×10^−37^).

**Figure 1.**
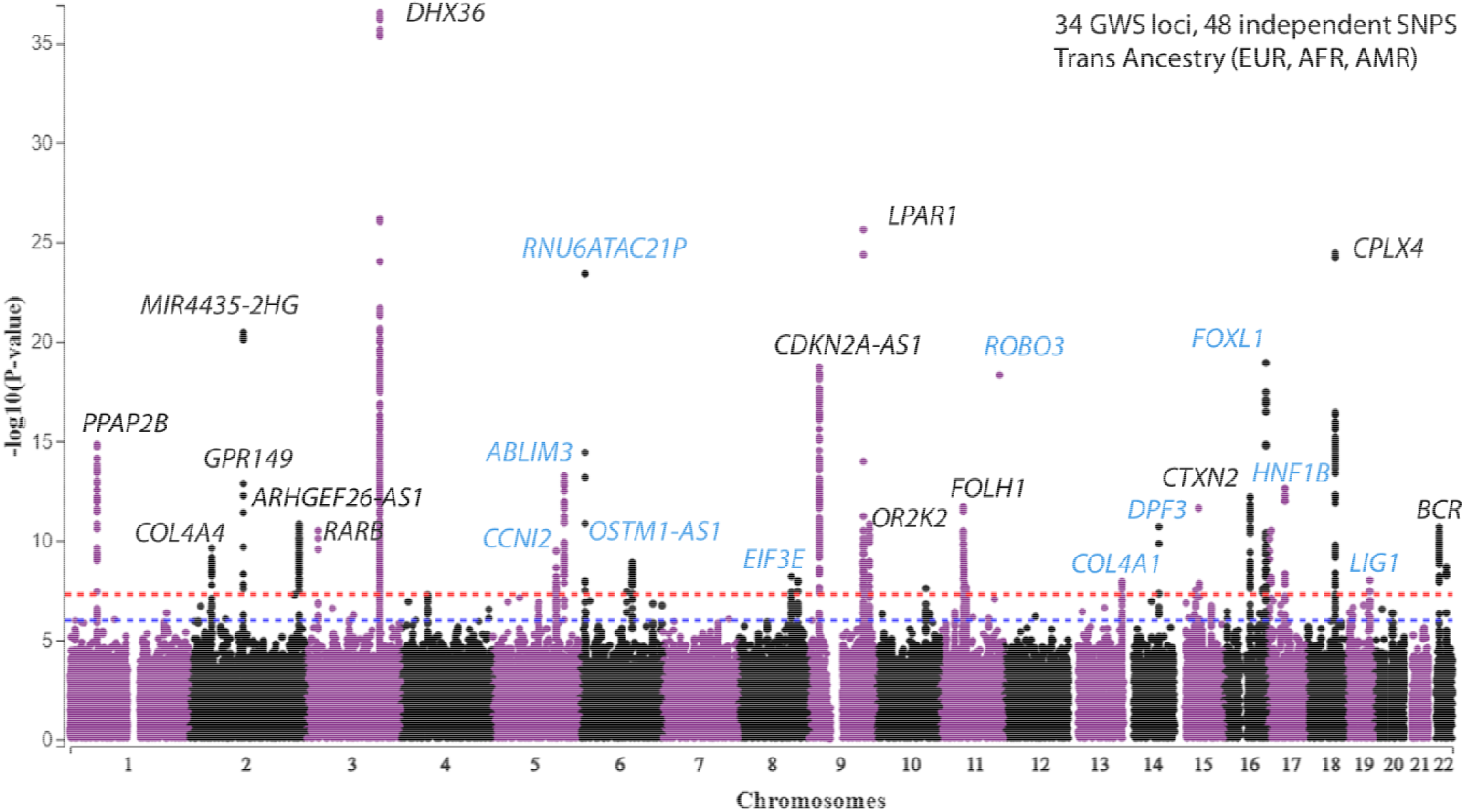
Trans-ancestry meta-analysis Manhattan Plot. Lead loci not identified in the European GWAS are highlighted.

**Table 3.**
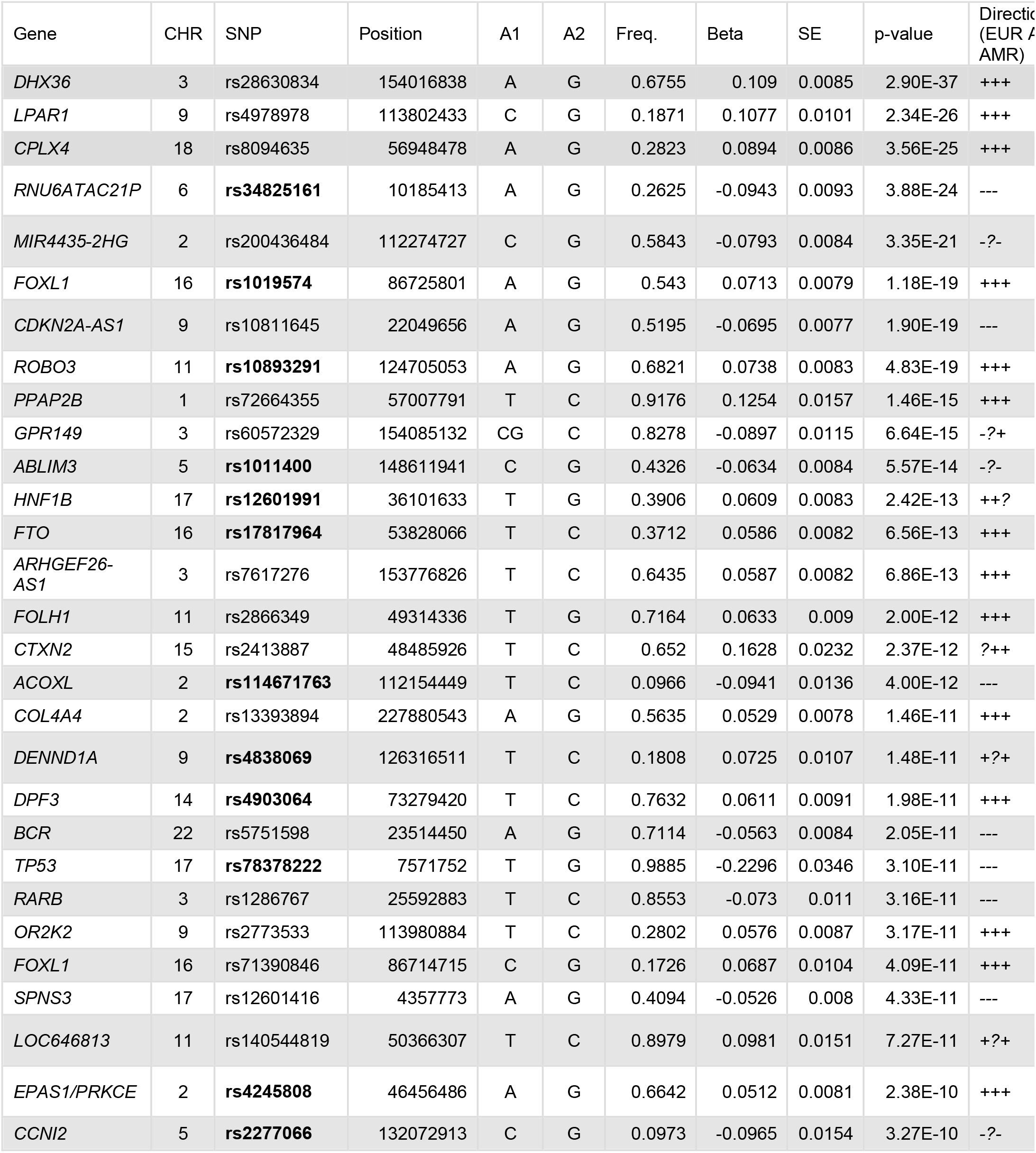
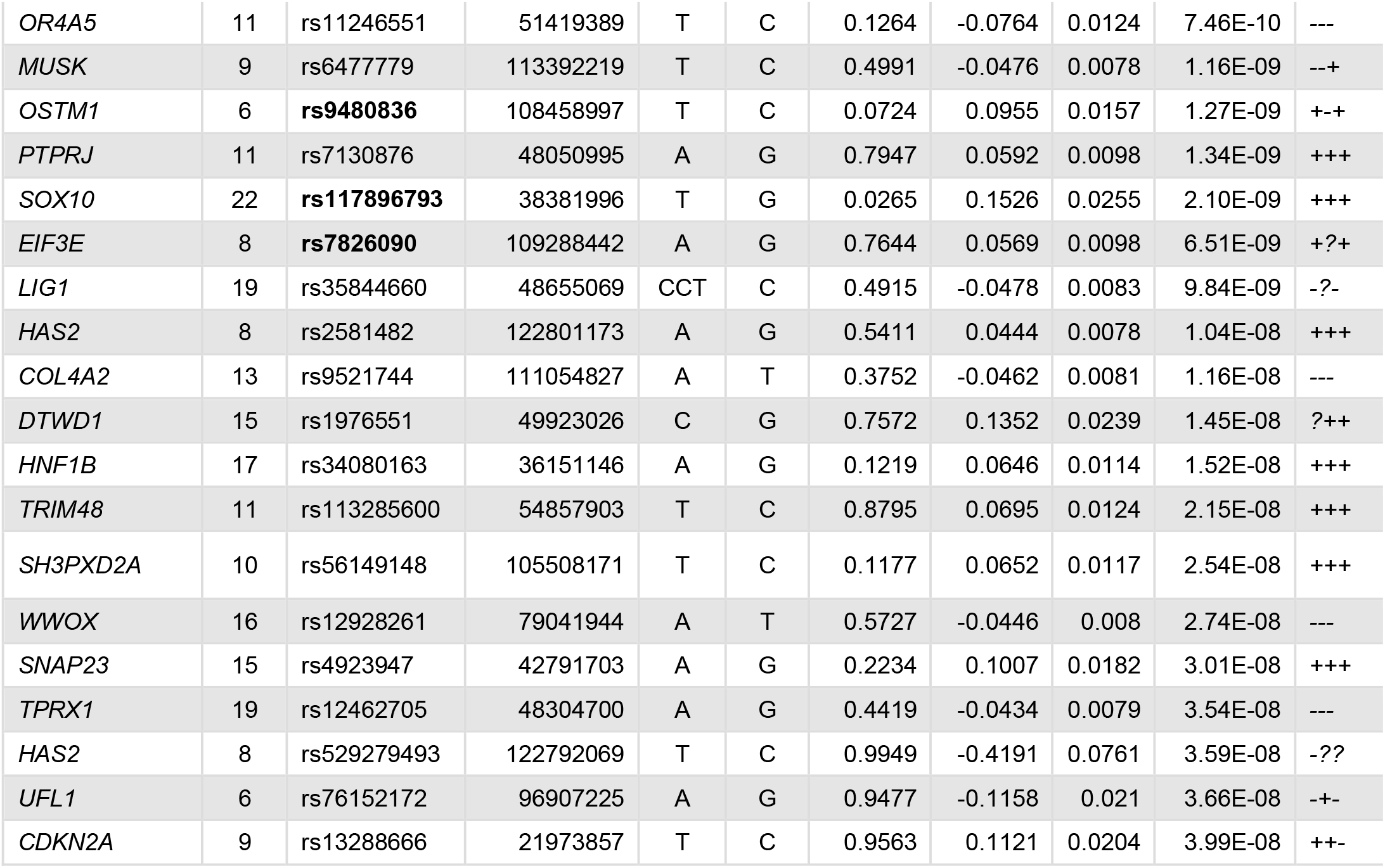
GWS variants identified in trans-ancestry meta. **Bolded variants** indicate cases where an identical variant was identified in the EUR only GWAS (15/34).

The only variant that was GWS in more than one population was *RNU6ATAC21P*\*rs34825161 (RNA, U6atac Small Nuclear 21, Pseudogene), significant in EUR (p=2.15×10^−18^), AFR (p=1.84×10^−8^), and also the trans-ancestry meta (p=3.88×10^−24^). The other variants identified in AFR and AMR may be population-specific.

### Heritability and Genetic correlations

The LDSC analysis showed significant heritability for EUR ERM (h^2^=0.050±0.006; p=3.41×10^−15^) and demonstrated significant positive genetic correlation (r_g_) between ERM and disorders of the vitreous body (r_g_=0.25±0.087; p=0.39×10^−2^), and depression (r_g_=0.14±0.039; p=0.04×10^−2^), whereas the genetic correlations between ERM and glaucoma and PTSD were not significant (rg=0.007±0.063, p=0.92; r_g_=0.080±0.057; p=0.16). The Manhattan plot for the GWAS on “disorders of the vitreous body” is provided in Figure S4. We also observed significant heritability in AFR (h^2^=0.064±0.031; p=0.039) but we obtained a non-significant heritability for AMR (h^2^=0.021±0.050; p=0.67). Due to lack of power and comparators, it was not possible to examine genetic correlations in these latter two populations.

### Cross-ancestry Fine-mapping for potential causal variants

We performed fine-mapping to identify potential causal variants in 33 loci identified by the cross-ancestry meta-analysis (Table S1). The median number of SNPs in the credible sets was 7. Six credible sets contain only a single variant with PIP ≥99% which could be targets for follow-up studies, including rs114671763, rs34825161, rs76152172, rs6477779, rs8094635 and rs117896793. There are 18 credible sets with ≤5 variants.

### Associated variants’ influence on gene expression

Two lead variants from the EUR GWAS were significantly associated with gene expression changes in the eye, rs1011400 and rs4245808 (Figure 2). Rs1011400 was significantly associated with changes in antisense RNA transcript *AC012613*.*2* in both macula (p=5.43×10^−16^, FDR=2.78×10^−12^) and non-macula retinal tissue (p=6.48×10^−12^, FDR=1.75×10^−8^). Rs1011400 was also significantly associated with changes in macula *ABLIM3* (p=1.22×10^−4^, FDR=0.036) expression. Rs4245808 was significantly associated with non-macular retinal pigment epithelium expression of *AC073283*.*1* (p=2.86×10^−5^, FDR=0.017). Three additional GWS proxy variants were associated with eye gene expression changes: rs2416908 (proxy for rs4838069) was associated with expression changes in *DENND1A* (the gene encoding DENN Domain Containing 1A) in macular retinal pigment epithelium (p=6.24×10^−5^, FDR=0.036), rs803141 (proxy for rs2277066) with *CCNI2* (encoding Cyclin I Family Member 2; p=1.14×10^−8^, FDR=2.26×10^−5^) and *SEPT8* (encoding Septin 8; p=3.00×10^−5^, FDR=0.018) in non-macular retinal pigment epithelium, and rs9438 (proxy for rs9823832) with *DHX36* in the macula (p=2.80×10^−5^, FDR=0.011).

**Figure 2.**
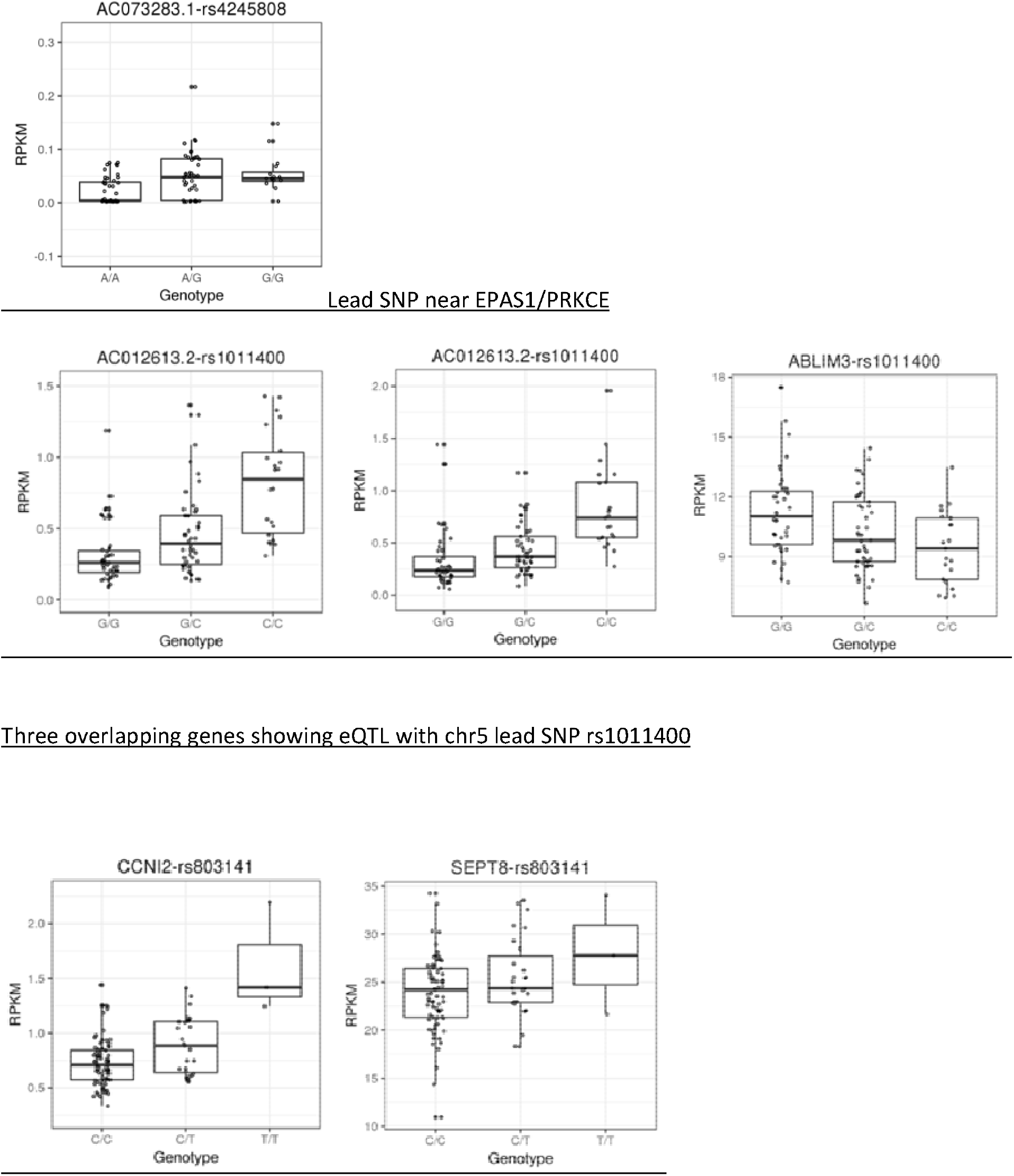
eQTL results. Expression QTLs were investigated in an eye tissue specific database. Lead SNPs were first looked up for each locus, with 2 significant results after multiple testing correction (FDR<0.05): rs4245808 and rs1011400. GWS proxy SNPs were selected when results were not available for the lead SNP, with three additional SNPs showing significant eQTL (FDR<0.05): rs803141, rs2416908, and rs9438. Genotype is on the X axis and Reads Per Kilobase Million (RPKM) is on the Y axis. Scale is variable and relative to the transcript depicted in each figure.

### Transcriptome-Wide Association Study (TWAS) and Fine-mapping

TWAS identified 28 independent associated genes (Table S2). The most significant gene was *DDX50P2* in testis (p=0.92×10^−24^), a pseudogene, followed by *CDKN2A* (p=0.29×10^−17^) in the brain cortex.

For TWAS fine-mapping, we obtained 12 genes with posterior inclusion probability (PIP) ≥ 0.9, and 7 more genes with PIP ≥ 0.7 (Table S3). Four genes had the highest PIP value (PIP=1): *CRIPT, MUSK, DDX50P2*, and *MALT1*.

### MAGMA gene-based and gene set analyses

MAGMA gene-based analysis provided a list of 25 significant genes which survived Bonferroni correction (p<2.62×10^−6^); these were provided as input for the gene set enrichment analysis (Table S4).

We obtained one significant term, the curated gene set of vitamin C in brain (Beta=1.296, STD=0.027, SE=0.282, p=2.12×10^−6^, P_bon_ =0.033).

### Functional Enrichment Analysis

There were significant enrichment results with six GO pathways (cyclin-dependent protein serine/threonine kinase inhibitor activity, collagen type IV trimer, networking-forming collagen trimer, collagen network, basement membrane collagen trimer, and complex of collagen trimers), and two significant protein complexes (CycD–Cdk4 and IL4-IL4R) (Table S4).

## Discussion

In this first well-powered GWAS of ERM conducted in the MVP, we identified 31 independent GWS risk loci in EUR participants, 3 independent loci in AFR, and 2 in AMR. In the trans-ancestry meta-analysis, 48 independent risk loci were identified. This trait is very important clinically – it is the most common retinal disease in adults^27^ -- and it causes substantial morbidity.

The most strongly supported variant in EUR was rs9823832 (p=9.06×10^−37^), at *DHX36* (DEAH-Box Helicase 36). In the trans-ancestry meta-analysis, the most strongly supported variant also maps to *DHX36* – a different variant at the same locus, rs28630834 (p=2.90×10^−37^). We did not identify any previous association of this locus to an ocular phenotype, but its relevance is supported by follow-up analysis that found that a GWS SNP in this locus (rs9438, p=6.38×10^−13^) alters expression of *DHX36* in macula (Figure 2). Considering some of the other associated variants of immediate biological interest in EUR, the protein product of *CPLX4* (rs7244528, p=2.84×10^−22^) plays a role in photoreceptor ribbon synapses^28,29^. *ROBO3*, to which variant rs10893291 maps (p=6.12×10^−17^), encodes Roundabout Guidance Receptor 3, involved in axonal navigation^30^. Rs1011400 maps to *ABLIM3* (p=2.87×10^−14^), which encodes an actin-binding protein related to nervous system development, previously reported to be associated to macular thickness^31^. We noted that variation at this locus alters expression in the macula of *ABLIM3* and *AC012613*.*2. FTO*\*rs17817964 (p=3.04×10^−11^) represents a locus well known for associations to body mass index-related traits. *RARB*\*rs57281891 (p=1.13×10^−10^) maps to the locus that encodes Retinoic Acid Receptor Beta, which binds retinoic acid. This locus has been previously associated to optic disc area^32^. Rs10120688 (*CDKN2B-AS1*) has been associated to primary open-angle glaucoma (POAG)^33^. *LPAR1*, the second-most-strongly associated locus both in EUR (p=1.81×10^−26^) and in the trans-ancestry meta-analysis (p=2.34×10^−26^) (different SNPs) encodes lysophosphatidic acid receptor 1. The protein product is a G-protein coupled receptor with a range of biological functions implicated in several traits with variants associated to eosinophil count^34^, and central corneal thickness^35^.

Other associated variants are linked to a range of different biological functions, e.g. two independent variants map to *ACOXL* (Acyl-Coenzyme A Oxidase-Like Protein), (rs6731210, p=5.93×10^−18^; rs114671763, p=1.13×10^−11^; fine-mapping revealed it is a potential causal variant with PIP >0.99) – this locus was previously associated to chronic lymphocytic leukemia^36^.

In AMR, the most strongly associated locus (p=2.21×10^−9^) was *CHRM2*, which encodes Cholinergic Receptor Muscarinic 2. Atropine, a mydriatic, is an agonist at the protein product of *CHRM2*, as is carbamoylcholine, an anti-glaucoma agent. Different variants at this locus have been shown to be associated to resting heart rate response to recovery after exercise^37^ and to risk-taking behavior^38^, among other phenotypes. Considering the association with ERM and *CHRM2* observed in AMR as well as *CDKN2B-AS1* (previously POAG-associated) in EUR, we explored the relationship of ERM and glaucoma further, but the genetic correlation between ERM and glaucoma was not significant; clinically, these traits (ERM and glaucoma) are not considered to be associated. LDSC was feasible only in EUR, and it is possible that this lack of a significant genetic correlation between these traits reflects an AMR-specific genetic relationship -- the lead *CHRM2* variant was monomorphic in EUR and not significant in AFR. However, there were significant genetic correlations between ERM and two other traits evaluated, depression and disorders of the vitreous (the latter from a FinnGen analysis). Visual impairment has been reported previously to be associated with increases in depression^39,40^. ERM is related to vitreous pathology, and risk is increased by vitreous detachment^27^.

The trans-ancestry meta-analysis identified 48 GWS variants, many not observed in any of the population-specific analyses. In numerous cases, the same gene was implicated but with a different lead variant, supporting shared biology between populations (Table 3). The only variant that was GWS in more than one population was *RNU6ATAC21P*\*rs34825161 (RNA, U6atac Small Nuclear 21, Pseudogene), significant in EUR, AFR, and the trans-ancestry meta-analysis. The other variants identified in AFR and AMR were not GWS in the meta-analysis, and these associations may be population-specific.

*CRIPT*, one of the genes identified by TWAS, encodes the Cysteine-Rich PDZ-Binding Protein, and high myopia is among several clinical traits associated with variants at this locus in a patient with an exon 1 deletion^41^. Another TWAS-associated gene, *CDKN2B-AS1*, which encodes Cyclin Dependent Kinase Inhibitor 2A, is associated with POAG, as noted above, and with several melanoma subtypes^42^. The protein product of *MUSK*, Muscle Associated Receptor Tyrosine Kinase, has been implicated in clustering of postsynaptic acetylcholine receptors in neuromuscular junction^43^ – interesting in the context of the *CHRM2* (Cholinergic Receptor Muscarinic 2) SNP-based association discussed above.

Pathway enrichment analysis using two different approaches revealed significant findings for collagen and for the collagen-adjacent vitamin C pathway. Vitamin C (ascorbic acid) is necessary for the hydroxylation of the proline residue in collagen, necessary to stabilize collagen fibers ^44^; MAGMA gene set enrichment analysis implicated vitamin C brain pathways. Collagen is a major constituent of epiretinal membranes, varying depending on the kind of membrane^45^ and a proteomic study has demonstrated that some collagens are upregulated in ERM^46^. Collagen type I and type IV are the two primary collagens founded in corneal and lens tissues; ^47^ g:profiler implicated collagen type IV trimer, network-forming collagen trimer, collagen network, basement membrane collagen trimer, and complex of collagen trimer (Table S4).

Above we highlight biology mostly consistent with prior knowledge, but our results also implicate more peripherally-connected biology. *FOXL1*, for example, encodes forkhead box L1. The same variant at this locus (rs1019574) is strongly associated to ERM in both EUR and the trans-population meta-analysis. Prior associations at this locus relate to, for example, bone mineral density^48^ and height^49^. It is also involved in eye development^50^, but may implicate a novel mechanism in the context of ERM.

Non-European populations are generally understudied with respect to GWAS. Since many genetic findings are at least to some extent population-specific, this is a major limitation that needs to be addressed by the research field as a whole^51^. For personalized medicine to become an effective strategy, population-specific polygenic risk scores will generally need to be available for each clinical population where a need for risk prediction is anticipated. This study demonstrates the strength of the MVP sample in addressing this issue, as we were able to study reasonable sample sizes of three major populations. Our findings contain a suggestion of some population-specific biology for ERM.

Our work has several limitations. ERM is diagnosed reliably via optical coherence tomography^52^ (OCT). Diagnoses recorded in the electronic health record (EHR) may reflect clinical or OCT diagnosis, but with large sample sizes signal overwhelms noise, tending to compensate for diagnosis imprecision. It is a limitation that we relied on EHR-based diagnosis and might have included secondary ERM cases. In relying on chart diagnosis that reflect the skills of a wide range of individual clinicians, it is likely that we included some false-positive amongst the cases with non-ERM pathology. However EHR diagnoses have been used widely and with great success previously for diagnosis of complex genetic traits (such as, for example, major depressive disorder^53^ and opioid use disorder^54^ specifically in the MVP), and the convergence of the identified risk loci identified with what is known about the pathophysiology of the disorder, supports that they are likely to be relevant to ERM. A further limitation is the mostly-male composition of the MVP sample; there may be sex-specific risk factors that we could not evaluate.

We identified GWS-significant associations to ERM in EUR, AFR, and AMR each taken individually, and many more GWS-associations in the trans-ancestry meta-analysis. Results from pathway enrichment analyses were consistent with known ERM pathophysiology. Availability of these genomewide results will enable the creation of polygenic risk scores (PRS) expected to be predictive of genetic risk for ERM. These findings should lead to improved understanding of the pathophysiology of ERM and, ideally, identifications of new targets for nonsurgical treatment interventions.

## Data Availability

Data will be made available through dbGAP upon publication of the article.
https://www.ncbi.nlm.nih.gov/projects/gap/cgi-bin/study.cgi?study_id=phs001672.v9.p1

## Acknowledgments

This research is based on data from the Million Veteran Program (MVP), Office of Research and Development, Veterans Health Administration, and was supported by the MVP and the Veterans Affairs Cooperative Studies Program study No. 575B. This work was also supported funding from the Department of Veterans Affairs Office of Research and Development grants I01CX001849 – MVP025. D.F.L. was supported by a was supported by an NARSAD Young Investigator Grant from the Brain & Behavior Research Foundation and a Career Development Award CDA-2 from the Veterans Affairs Office of Research and Development (1IK2BX005058-01A2) and is Aimee Mann Fellow of Psychiatric Genetics. This publication does not represent the views of the Department of Veterans Affairs or the United States Government. We also acknowledge the participants and investigators of the FinnGen study.

## Author contributions

JG conceived the study, drafted the article, and supervised the work. DL and MG analyzed the data and contributed to drafting the article. HZ and KA contributed to the data analysis. KH contributed to the phenotyping effort in the MVP sample. JG, MBS, and JMG provided administrative support. Additionally, all authors (DL, MG, KH, HZ, KA, JMG, DE, and MBS) reviewed the article and provided critical comments.

## Data Availability

Summary statistics will be made available in dbGAP (accession number phs001672) by the Million Veteran Program following publication.

