## Supplemental Information, Tables, and Figures for "Genomewide association study of epiretinal membrane: discovery of significant risk loci in each of three American populations"

Supplementary Information

1. Supplementary methods

**Diagnosis definitions**

**MVP**

Diagnosis definition for epiretinal membrane (ERM) in MVP: ICD10 codes H35.371 - Puckering of macula, right eye; H35.372 - Puckering of macula, left eye; and H35.373 - Puckering of macula, bilateral; H35.379 - Puckering of macula, unspecified eye; and ICD9 code 362.56 - Macular puckering.

**FinnGen**

The definition of the trait “Diseases of the Vitreous Body” includes ICD-10 H43 Disorders of the vitreous body; specifically, H43.0 Vitreous prolapse excluding vitreous syndrome following cataract surgery (H59.0); H43.1 Vitreous hemorrhage; H43.2 Crystalline deposits in vitreous body; H43.3 Other vitreous opacities -Vitreous membranes and strands; and H43.8 Other disorders of vitreous body - Vitreous: degeneration, detachment, excluding: proliferative vitreo-retinopathy with retinal detachment (H33.4); and H43.9 Disorder of vitreous body, unspecified.

The definition of the trait “Glaucoma” includes codes ICD-10 H40 Glaucoma, and H42 Glaucoma in diseases classified elsewhere; specifically, H40.0 Glaucoma suspect; H40.1 Primary open-angle glaucoma; H40.2 Primary angle-closure glaucoma; H40.3 Glaucoma secondary to eye trauma; H40.4 Glaucoma secondary to eye inflammation; H40.5 Glaucoma secondary to other eye disorders; H40.6 Glaucoma secondary to drugs; H40.8 Other glaucoma; H40.9 Glaucoma, unspecified, H42.0 Glaucoma in endocrine, nutritional and metabolic diseases; H42.8 Glaucoma in other diseases classified elsewhere.

For “diseases of the vitreous body,” there were 10,849 cases (3969 males and 6880 females) and 297,889 controls. For glaucoma, there were 13,614 cases (5759 males and 7855 females) and 295,540 controls.

**MVP Analysis**

*Genotyping, Imputation, Quality Control (QC), and GWAS details:*

Indels and complex variants were imputed independently using the 1000 Genomes (1KG) phase 3 panel and merged in an approach similar to that employed by the UK Biobank. Designation of ancestries was based on genetic assignment with comparison to 1KG reference panels^1^.

MVP GWAS was conducted using logistic regression in PLINK 2.0 using the first 10 ancestry principal component (PCs), sex, and age as covariates. SNPs with minor allele frequency (MAF) <0.1%, genotype missingness >20%, Hardy-Weinberg equilibrium p<5x10^-7^ , and all but one individual from every kinship coefficient pairing ≥0.0884 (≥2^rd^-degree relation) were excluded randomly. Trans-ancestry meta-≤analysis was performed in METAL using inverse variance weighting. SNPs were assigned to genes in Tables 2 and 3 based on proximity (within 100 kb).

**FinnGen analysis (as done by Finngen)**

Genotype data were generated using two different arrays (one Illumina and one Affymetrix). The available summary statistics were calculated from genotype data excluding individuals with ambiguous sex and genotype missingness >0.05. GWASs were run using Regenie version 2.0.2 ^2^, including the first 10 PCs, sex, age, and genotyping batch as covariates. SNPs with missingness >0.02, Hardy-Weinberg equilibrium p<10^-6^ and low minor allele count ≤2 were excluded. Eagle 2.3.5 ^3^ was used for pre-phasing, with conditioning haplotypes set to 20.000 and the other default parameters, followed by genotype imputation which was done with the population-specific SISu v3 reference panel (<https://github.com/FINNGEN/finngen-documentation/blob/8a24390c151773efba74d97af7209a3acde32fa9/methods/genotype-imputation/sisu-reference-panel.md>) with Beagle 4.1 ^4^.

**Cross-ancestry Fine Mapping**

For each of the 48 lead variants identified in cross-ancestry meta-analyses, we performed clumping using PLINK (within 500 kb and LD r^2^>0.1) in EUR, AFR, and AMR summary data separately to obtain 3 sets of SNPs (p<0.05) for fine-mapping; corresponding LD reference panels from 1000 Genomes Project were used. For loci with lead SNPs not present in the EUR SNP set, or with limited numbers of variants which could not have convergent results, we did not perform fine-mapping. After these exclusions, 33 loci were analyzed. We assumed initially that each locus contained only one causal variant by default, increased to three at maximum if the analysis unable to converge. Credible set was defined as plausible causal variants with accumulated posterior inclusion probability (PIP) >99%.

*Transcriptome-Wide Association Study (TWAS) and Fine-mapping*

We used GTEx_v8^5; 6^ multi tissue expression for the EUR samples (<https://www.mancusolab.com/>) which are available for 49 tissues. For the linkage disequilibrium reference data, we used the 1000 Genomes LD reference (<https://data>.broadinstitute.org/alkesgroup/FUSION/LDREF.tar.bz2). We initially identified associated genes, then post-processed these results to distinguish the conditionally independent genes, because of the possibility of multiple associated features at some loci. Thus, after discarding the conditionally non-significant genes, for each gene we kept the tissue where the most significant TWAS p-value was provided.

To fine-map TWAS statistics with respect to genomic risk regions, we used FOCUS^7^ for the EUR statistics. GTEx_v7 weights were combined with Metabolic Syndrome in Men Study (METSIM), Netherlands Twins Registry (NTR), Young Finns Study (YFS), and CommonMind Consortium (CMC) weights. LD scores were obtained from 1000 Genome phase 3.

*MAGMA gene-based and gene set analyses*

Gene-based analysis calculates p-value for protein-coding genes by mapping SNPs to genes if SNPs are located within them ^8^. Gene-set analysis was performed for curated gene-sets and gene ontology (GO) terms obtained from MsigDB, considering as significant those results with Bonferroni corrected p-value (P_bon_)<0.05 (p<2.62x10^-6^, reflecting correction for 19,106 genes tested). The g:Profiler toolset ^9^ was used to perform functional enrichment analysis including GO, biological pathways, regulatory motifs in DNA, tissue specificity and protein complexes databases, and human disease phenotypes. It has a web interface allowing the direct upload of a list of genes, with data that can belong to any organism. We provided a list of 25 genes obtained from MAGMA gene-based analysis of the cross-ancestry meta-analysis.

2. Supplementary figures

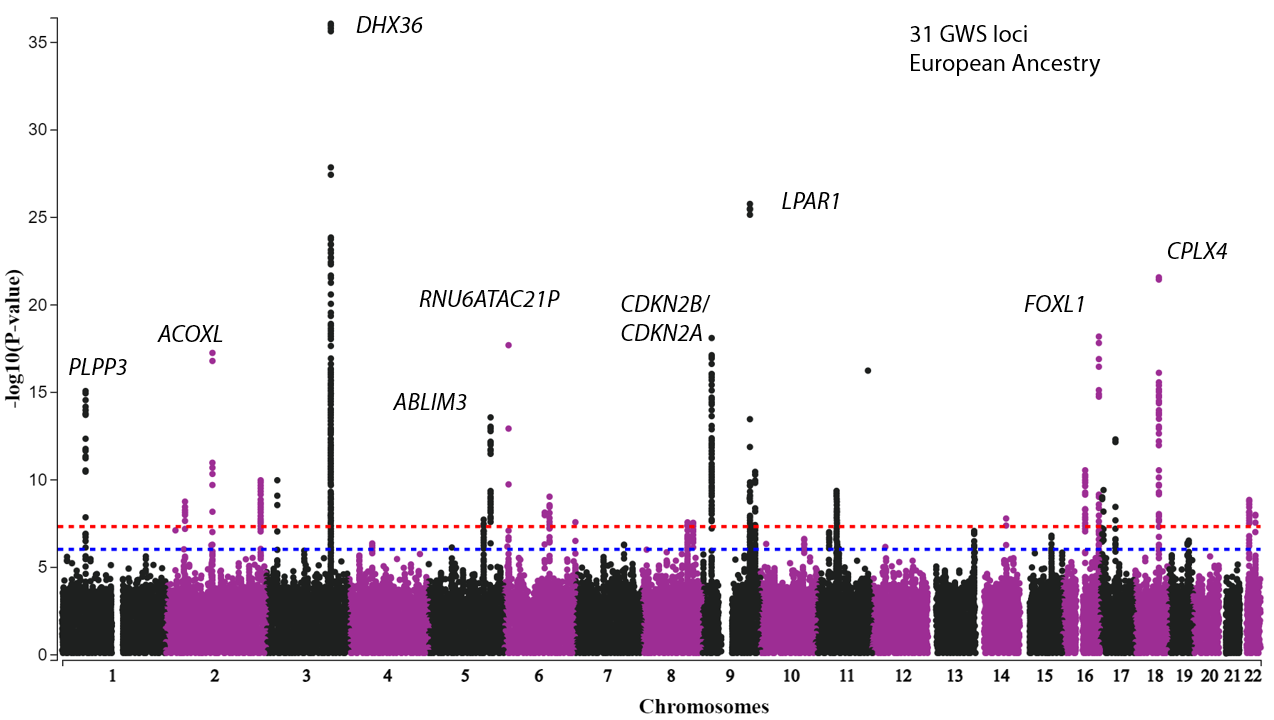

Figure S1. ERM GWAS in EUR. 31 independent genomewide-significant risk loci were identified.

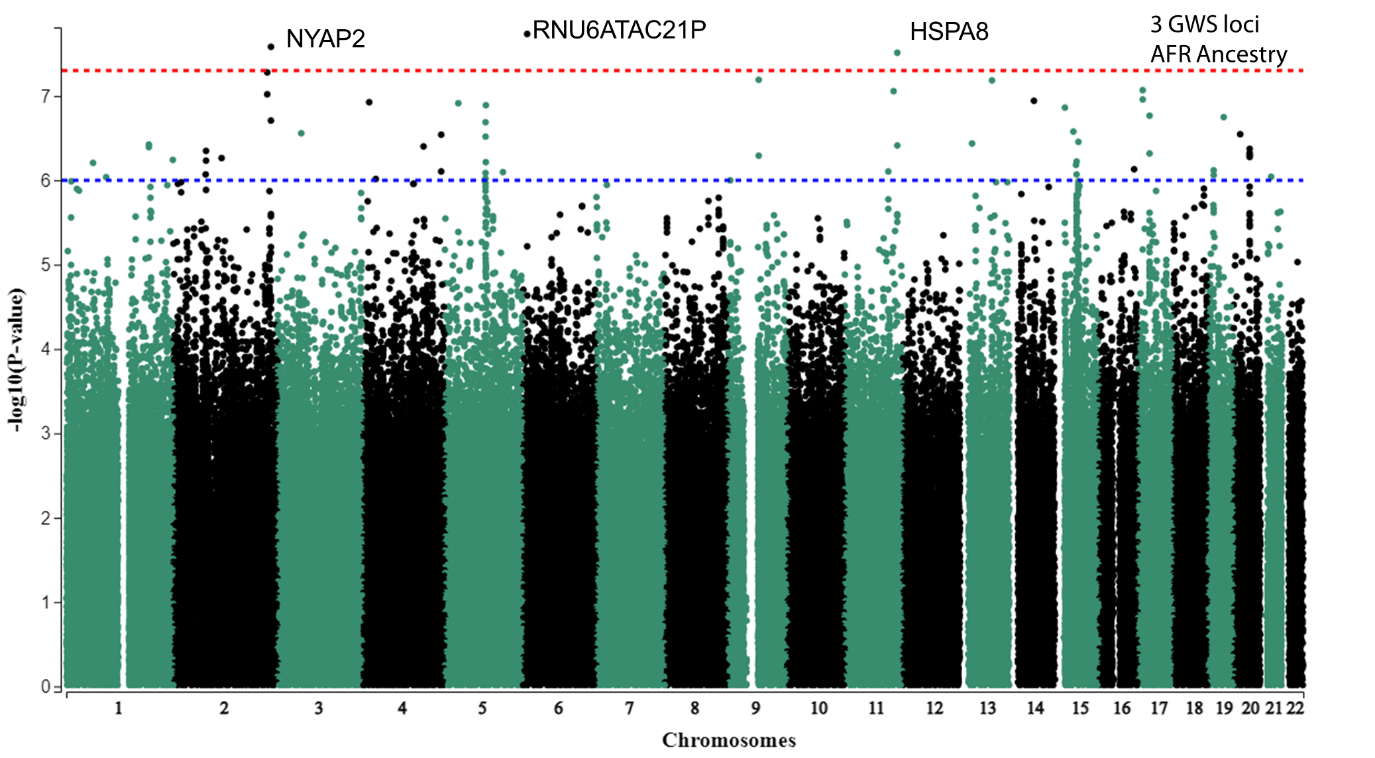

Figure S2. ERM GWAS in AFR. 3 independent genomewide-significant risk loci were identified.

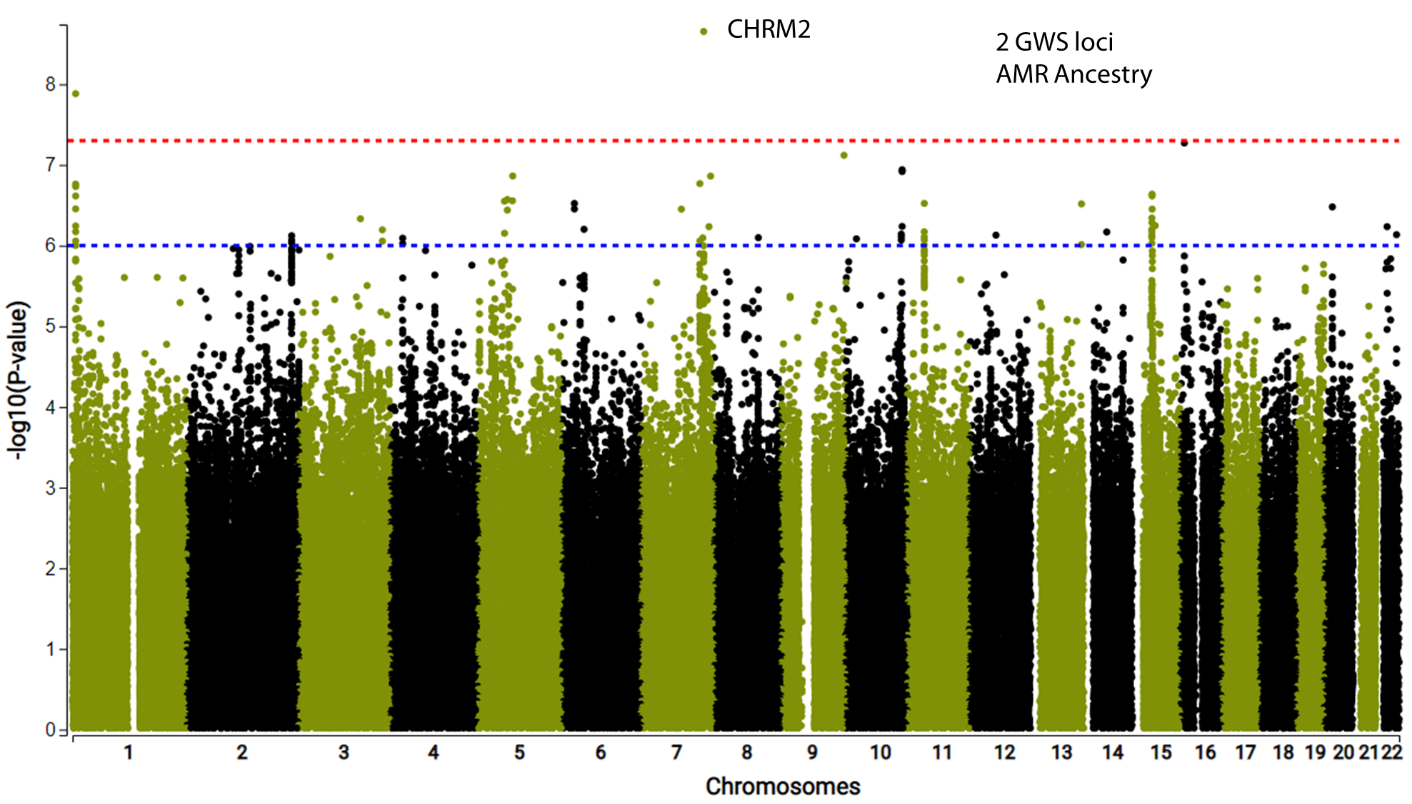
 Figure S3. ERM GWAS in AMR. One independent genomewide-significant risk loci was identified.

Figure S4.

Manhattan plot: disorders of vitreous body (Finngen data)

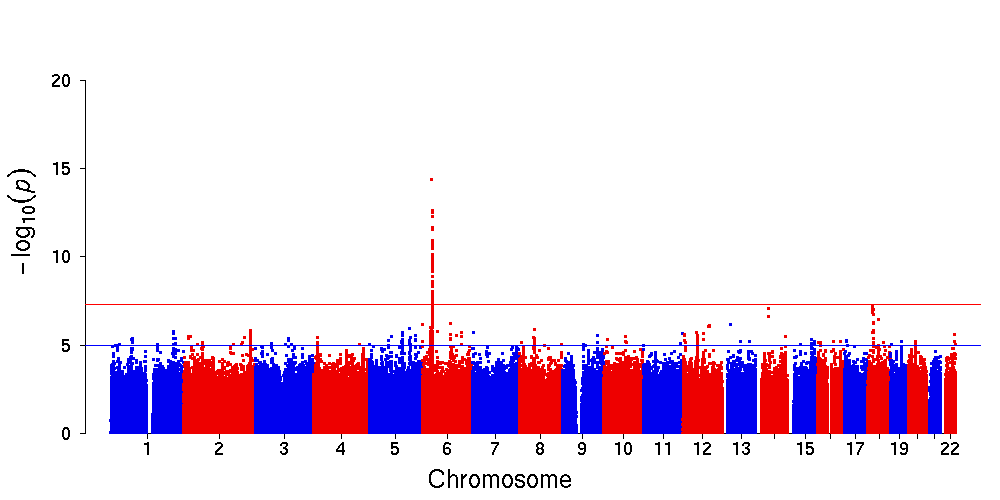

3. Supplementary tables

| **Chr** | **rsID** | **pos** | **Populations** | **Clumped region** | **#SNPs** | **CS_length** | **BestPIP_pos** | **BestPIP** | **LeadSNP_PIP** |
| --- | --- | --- | --- | --- | --- | --- | --- | --- | --- |
| 2 | rs4245808 | 46456486 | EUR,AFR,AMR | 46373596:46456486 | 68 | 15 |  | 0.275 | 0.275 |
| 2 | rs114671763 | 112154449 | EUR,AFR,AMR | 111932275:112319781 | 10 | 1 |  | 0.999 | 0.999 |
| 2 | rs200436484 | 112274727 | EUR,AMR | 112253302:112327070 | 12 | 4 | chr2:112270407 | 0.345 | 0.128 |
| 2 | rs13393894 | 227880543 | EUR,AFR,AMR | 227698797:227955447 | 43 | 16 |  | 0.220 | 0.220 |
| 3 | rs1286767 | 25592883 | EUR,AFR,AMR | 25554363:25603749 | 25 | 4 | chr3:25581512 | 0.329 | 0.289 |
| 5 | rs2277066 | 132072913 | EUR,AMR | 131995843:132179445 | 63 | 12 |  | 0.707 | 0.707 |
| 5 | rs1011400 | 148611941 | EUR,AMR | 148549111:148659664 | 31 | 8 |  | 0.556 | 0.556 |
| 6 | rs34825161 | 10185413 | EUR,AFR,AMR | 10149706:10235326 | 20 | 1 |  | 1.000 | 1.000 |
| 6 | rs76152172 | 96907225 | EUR,AFR,AMR | 96907225:97278873 | 9 | 1 |  | 0.996 | 0.996 |
| 6 | rs9480836 | 108458997 | EUR,AFR,AMR | 108425822:108462792 | 12 | 8 |  | 0.273 | 0.273 |
| 8 | rs529279493 | 122792069 | EUR | 122499564:122826201 | 4 | 4 | chr8:122597340 chr8:122826201 | 0.250 0.250 | 0.250 |
| 8 | rs2581482 | 122801173 | EUR,AFR,AMR | 122631762:122837425 | 37 | 11 |  | 0.853 | 0.853 |
| 9 | rs13288666 | 21973857 | EUR,AFR,AMR | 21956492:22022990 | 7 | 2 | chr9:21973857 chr9:21956492 | 0.50 0.50 | 0.500 |
| 9 | rs6477779 | 113392219 | EUR,AFR,AMR | 112893236:113547476 | 43 | 1 |  | 0.999 | 0.999 |
| 9 | rs4978978 | 113802433 | EUR,AFR,AMR | 113719411:113841657 | 19 | 4 | chr9:113792997 | 0.400 | 0.375 |
| 10 | rs56149148 | 105508171 | EUR,AFR,AMR | 105480149:105731546 | 44 | 7 |  | 0.907 | 0.907 |
| 11 | rs11246551 | 51419389 | EUR,AFR,AMR | 51126759:51580744 | 40 | 19 | chr11:51252962 | 0.339 | 0.000 |
| 11 | rs113285600 | 54857903 | EUR,AFR,AMR | 54824098:55353966 | 44 | 22 |  | 0.240 | 0.240 |
| 13 | rs9521744 | 111054827 | EUR,AFR,AMR | 111016886:111114554 | 54 | 18 | chr13:111054826 | 0.331 | NA |
| 14 | rs4903064 | 73279420 | EUR,AFR,AMR | 73245571:73522108 | 29 | 2 |  | 0.870 | 0.870 |
| 16 | rs17817964 | 53828066 | EUR,AFR,AMR | 53573178:53845487 | 40 | 6 |  | 0.636 | 0.636 |
| 16 | rs12928261 | 79041944 | EUR,AFR,AMR | 79037971:79046467 | 10 | 2 |  | 0.601 | 0.601 |
| 16 | rs71390846 | 86714715 | EUR,AFR,AMR | 86623034:86719771 | 22 | 3 |  | 0.795 | 0.795 |
| 16 | rs1019574 | 86725801 | EUR,AFR,AMR | 86667409:86731223 | 64 | 4 |  | 0.910 | 0.910 |
| 17 | rs12601416 | 4357773 | EUR,AFR,AMR | 4325323:4382729 | 38 | 4 |  | 0.623 | 0.623 |
| 17 | rs78378222 | 7571752 | EUR,AFR,AMR | 7316030:7578671 | 8 | 3 | chr17:7578671 | 0.986 | NA |
| 17 | rs12601991 | 36101633 | EUR,AFR | 35632979:36208392 | 26 | 2 |  | 0.704 | 0.704 |
| 17 | rs34080163 | 36151146 | EUR,AFR,AMR | 36135143:36245376 | 15 | 6 |  | 0.664 | 0.664 |
| 18 | rs8094635 | 56948478 | EUR,AFR,AMR | 56899666:57128486 | 58 | 1 |  | 1.000 | 1.000 |
| 19 | rs12462705 | 48304700 | EUR,AFR,AMR | 48243036:48352054 | 44 | 7 |  | 0.894 | 0.894 |
| 19 | rs35844660 | 48655069 | EUR,AMR | 48613544:48688683 | 45 | 18 |  | 0.273 | 0.273 |
| 22 | rs5751598 | 23514450 | EUR,AFR,AMR | 23319952:23624093 | 73 | 22 |  | 0.272 | 0.272 |
| 22 | rs117896793 | 38381996 | EUR,AFR,AMR | 38163777:38620179 | 12 | 1 |  | 0.996 | 0.996 |

Table S1. Fine-mapping for regions with independent variants in cross-ancestry meta-analysis. 33 regions were analyzed. #SNPs are the number of SNPs included in the analysis; CS_length, credible set length; PIP, posterior inclusion probability.

| ID | TWAS.Z | TWAS.P | hgnc_symbol | tissue |
| --- | --- | --- | --- | --- |
| ENSG00000240048 | -10 | 9.2e-25 | *DDX50P2* | Testis |
| ENSG00000147889 | -8.7 | 2.9e-18 | *CDKN2A* | Brain_Cortex |
| ENSG00000114790 | -8.6 | 1.1e-17 | *ARHGEF26* | Artery_Aorta |
| ENSG00000147883 | 8.5 | 1.7e-17 | *CDKN2B* | Muscle_Skeletal |
| ENSG00000174953 | 8.3 | 1.3e-16 | *DHX36* | Pancreas |
| ENSG00000162407 | -8 | 9e-16 | *PLPP3* | Brain_Cortex |
| ENSG00000173210 | 7.4 | 1.1e-13 | *ABLIM3* | Brain_Anterior_cingulate_cortex_BA24 |
| ENSG00000157510 | -7.4 | 1.8e-13 | *AFAP1L1* | Cells_Cultured_fibroblasts |
| ENSG00000238755 | -7.4 | 1.9e-13 | *LINC02006* | Esophagus_Mucosa |
| ENSG00000253406 | -7.4 | 1.9e-13 |  | Adipose_Subcutaneous |
| ENSG00000255199 | -6.7 | 1.8e-11 | *GTF2IP11* | Testis |
| ENSG00000243069 | 6.4 | 1.2e-10 | *ARHGEF26-AS1* | Esophagus_Gastroesophageal_Junction |
| ENSG00000261175 | -6.3 | 2.2e-10 | *LINC02188* | Thyroid |
| ENSG00000144468 | 6.1 | 1.4e-09 | *RHBDD1* | Artery_Tibial |
| ENSG00000168955 | 6 | 1.9e-09 | *TM4SF20* | Breast_Mammary_Tissue |
| ENSG00000274478 | 5.8 | 6.8e-09 |  | Artery_Aorta |
| ENSG00000119522 | 5.7 | 1.2e-08 | *DENND1A* | Heart_Atrial_Appendage |
| ENSG00000149177 | -5.7 | 1.2e-08 | *PTPRJ* | Colon_Transverse |
| ENSG00000081052 | -5.7 | 1.3e-08 | *COL4A4* | Adipose_Visceral_Omentum |
| ENSG00000086205 | -5.7 | 1.4e-08 | *FOLH1* | Skin_Not_Sun_Exposed_Suprapubic |
| ENSG00000156374 | 5.6 | 1.6e-08 | *PCGF6* | Pituitary |
| ENSG00000100218 | -5.6 | 1.7e-08 | *RSPH14* | Whole_Blood |
| ENSG00000255042 | -5.6 | 1.7e-08 | *SEPTIN7P11* | Pituitary |
| ENSG00000030066 | 5.6 | 1.9e-08 | *NUP160* | Whole_Blood |
| ENSG00000196666 | 5.6 | 1.9e-08 | *FAM180B* | Colon_Sigmoid |
| ENSG00000057294 | -5.6 | 2.2e-08 | *PKP2* | Whole_Blood |
| ENSG00000100228 | 5.6 | 2.4e-08 | *RAB36* | Adrenal_Gland |
| ENSG00000131437 | 5.5 | 3.6e-08 | *KIF3A* | Artery_Coronary |

Table S2. Independent associated loci from TWAS.

| mol_name | tissue | twas_z | pip |
| --- | --- | --- | --- |
| *CRIPT* | pituitary | 5.95 | 1 |
| *MUSK* | brain_caudate_basal_ganglia | -10.4 | 1 |
| *DDX50P2* | testis | -9.62 | 1 |
| *MALT1* | muscle_skeletal | 8.45 | 1 |
| *PPAP2B* | brain_cortex | -7.82 | 0.998 |
| *TXNDC5* | brain_cerebellar_hemisphere | 4.98 | 0.963 |
| *DENND1A* | esophagus_muscularis | 4.93 | 0.957 |
| *CRX* | testis | -4.95 | 0.943 |
| *RP11-574M7.2* | testis | 6.68 | 0.933 |
| *IRF8* | minor_salivary_gland | -4.56 | 0.929 |
| *PSMB3* | esophagus_mucosa | -5.65 | 0.919 |
| *FOXO1* | cells_transformed_fibroblasts | 4.75 | 0.9 |
| *SYCP2L* | brain_amygdala | 4.82 | 0.899 |
| *PKP2* | whole_blood | -4.66 | 0.866 |
| *ARMC2* | brain_putamen_basal_ganglia | -5.88 | 0.806 |
| *EXOC7* | minor_salivary_gland | -4.57 | 0.778 |
| *TBX3* | lung | -4.51 | 0.765 |
| *DPF3* | artery_tibial | -4.98 | 0.709 |
| *TRIM48* | skin_sun_exposed_lower_leg | 4.46 | 0.707 |

Table S3. Genes with PIP ≥ 0.7

| **g:profiler** | | **MAGMA Gene-Set Analysis** | | |
| --- | --- | --- | --- | --- |
| Term name | adjusted p | Gene Set | P | P bon |
| *cyclin-dependent protein serine/threonine kinase inhibitor activity* | 0.0243 | ***Curated gene sets:biocarta vit c brain pathway*** | 2.12E-06 | 0.033 |
| ***collagen type IV trimer*** | 0.0042 | *GO_bp:go_regulation of fc receptor mediated stimulatory signaling pathway* | 6.44E-06 | 0.100 |
| ***network-forming collagen trimer*** | 0.0072 | *GO_mf:go_syntaxin_binding* | 1.37E-05 | 0.212 |
| ***collagen network*** | 0.0072 | *GO_mf:go_cyclin dependent protein serine threonine kinase inhibitor activity* | 1.74E-05 | 0.269 |
| ***basement membrane collagen trimer*** | 0.0072 | *GO_cc:go_collagen_type_iv_trimer* | 2.74E-05 | 0.424 |
| ***complex of collagen trimers*** | 0.0456 | *Curated gene sets:reactome sumoylation of transcription factors* | 2.96E-05 | 0.459 |
| *IL4-IL4R complex* | 0.0497 | *GO_bp:go cellular response to insulin like growth factor stimulus* | 2.99E-05 | 0.462 |
| *p16-cyclin D2-CDK4 complex* | 0.0497 | *GO_mf:go_snare_binding* | 4.06E-05 | 0.629 |

Table S4. Eight pathway terms were significantly enriched following multiple testing correction in g:profiler (left), while only 1 pathway was significant in the MAGMA gene-set analysis (right) following correction (vitamin c pathway, defined by the GSEA resource as “vitamin C in the brain”; https://www.gsea-msigdb.org/gsea/msigdb/cards/BIOCARTA_VITCB_PATHWAY).  Bolding represents pathways discussed in greater detail within text. Pbon = Bonferroni corrected P value.
